# Impact of SARS-CoV-2 vaccination on systemic immune responses in people living with HIV

**DOI:** 10.1101/2022.04.08.22273605

**Authors:** Clara Bessen, Carlos Plaza-Sirvent, Jaydeep Bhat, Corinna Marheinecke, Doris Urlaub, Petra Bonowitz, Sandra Busse, Sabrina Schumann, Elena Vidal Blanco, Adriane Skaletz-Rorowski, Norbert H Brockmeyer, Oliver Overheu, Anke Reinacher-Schick, Simon Faissner, Carsten Watzl, Stephanie Pfaender, Anja Potthoff, Ingo Schmitz

## Abstract

The severe acute respiratory syndrome coronavirus 2 (SARS-CoV-2) causes coronavirus disease 2019 (COVID-19) and an ongoing global pandemic. Despite the development of vaccines, which protect healthy people from severe and life-threatening COVID-19, the immunological responses of people with secondary immunodeficiencies to SARS-CoV-2 mRNA vaccines are currently not well understood. Human Immunodeficiency Virus (HIV), causing acquired immunodeficiency syndrome (AIDS), targets CD4+ T helper (Th) cells that orchestrate the immune response. Anti-retroviral therapy suppresses HIV burden and restores Th cell numbers. Here, we investigated the humoral and cellular immune responses elicited by the BTN162b2 vaccine in a cohort of people living with HIV (PLWH), who receive anti-retroviral therapy. While antibody responses in PLWH increased progressively after the first and second vaccination compared to baseline, they were reduced compared to HIV negative study participants (controls). CD8+ T cells exhibited a general activated phenotype and increased effector and effector memory compartments. In contrast, CD4+ Th cell responses exhibited a vaccination-dependent increase and were comparable between PLWH and controls. In line with their reduced humoral response, the correlation between neutralizing antibodies and the CD4+ T cell response was decreased in PLWH compared to healthy controls. Interestingly, CD4+ T cell activation negatively correlated with the CD4 to CD8 ratio, indicating that low CD4 T cell numbers do not necessarily interfere with cellular immune responses. Taken together, our data demonstrate that COVID-19 mRNA vaccination in PLWH results in potent cellular immune responses, but the reduced antibody responses suggest that booster vaccination might be required for preventing disease.

## INTRODUCTION

Since early 2020, the world is suffering from a pandemic caused by severe acute respiratory syndrome coronavirus 2 (SARS-CoV-2, also known as 2019-nCoV) ^1^. Clinical manifestations of coronavirus disease 2019 (COVID-19), the disease caused by SARS-CoV-2, diverge from asymptomatic stage or mild influenza-like symptoms to death due to the acute respiratory failure. Most infected patients recover without the need for hospital care, but factors such as age or existence of comorbidities, like diabetes or immunodeficiency, determine the probability to develop severe COVID-19 ^2, 3^. SARS-CoV-2 infection elicits humoral responses, most importantly antibodies against viral proteins, as well as a wide variety of cellular responses ^4, 5, 6, 7, 8, 9, 10^. Among other measures to control the pandemic, vaccination strategies have been implemented to prevent severe COVID-19 ^11^. In contrast to traditional vaccination strategies, a novel technology of mRNA-based vaccines has been implemented. The novel mRNA-based vaccines deliver *in vitro* transcribed mRNA molecules encoding for a pathogen antigen, which are encapsulated in lipid nanoparticles ^12^. In case of COVID-19 mRNA-based vaccines, the SARS-CoV-2 spike protein, which binds to the human angiotensin-converting enzyme 2 (ACE2) receptor, is translated and presented by antigen-presenting cells, generating a specific immune response ^12, 13^. Besides safety and efficacy, studies demonstrated that these vaccines are able to generate a durable response ^14, 15, 16, 17, 18^. This immune response comprises the induction of germinal centers and production of vaccine-induced antibodies as well as the generation of spike-specific T cells ^17, 19, 20^. Nevertheless, due to a decline in the antibody titers over time and the rise of SARS-CoV-2 variants of concern, a vaccine booster dose is recommended to preserve the protective effect against COVID-19 ^21, 22, 23^.

Vaccine safety and efficacy are being stringently monitored during vaccine development and vaccination campaign. Still, the effectiveness of vaccine is not completely clear in patients with immunodeficiency such as human immunodeficiency virus (HIV) infection. Currently, it is poorly understood whether people living with HIV (PLWH) have a higher risk to develop severe COVID-19. In initial COVID-19 cohort studies, comparable clinical outcomes were found in the PLWH and HIV-negative individuals ^24, 25, 26, 27, 28, 29, 30^. Contrariwise, in other cohort studies, PLWH presented worse outcomes including higher rates of hospitalization and mortality ^31, 32, 33, 34, 35^. In addition, an observational study described lower IgG concentration and neutralizing antibody titers correlating with more cases of severe COVID-19 in PLWH ^36^. In contrast, a recent work investigating humoral and T cell specific-responses to the SARS-CoV-2 infection revealed that PLWH could mount a persistent immune response comparable to HIV-negative subjects ^37^. Additionally, the correlation analysis in the latter study suggested that deviations in the CD4 to CD8 ratio may result in a diminished ability to respond to SARS-CoV-2 infection in the PLWH ^37^. Taken together, the current data on SARS-CoV-2 infection and PLWH raises the question whether and how COVID-19 vaccines raise immune responses. Our present study aims to clarify whether immunity elicited by a COVID-19 mRNA vaccine in the PLWH is modulated in comparison to vaccination in HIV-negative people.

## MATERIALS AND METHODS

### Study design

The study was authorized by the local ethics committee of the Ruhr-University Bochum (21-7351 and 20-6953-bio). Study participants were recruited at the WIR - Walk in Ruhr, Department of Dermatology, Ruhr-University Bochum. We included healthy age-matched HIV-negative participants (controls; n=20) and PLWH (n=71). The written informed consent was obtained from all the patients. The clinical information of the study participants is presented in Table 1. Samples were collected immediately before the first vaccination (T0), at the day of second vaccination (T1) and within 4-6 weeks (T2) following the second vaccination against SARS-CoV-2.

**Table 1.** Clinical data of the cohort. Numbers (n) of people in the control and people living with HIV (PLWH) groups. Given are age, sex (F female; M male), HIV status, and anti-retroviral therapy (ART).

|  | **Control** | **PLWH** |
| --- | --- | --- |
| **n** | 20 | 71 |
| **Age (Mean ± SD)** | 39.4 ± 11.9 | 46.1 ± 10.9 |
| **Sex (F/M)** | 12/8 | 9/62 |
| **HIV** | - | + |
| **ART** | - | + |

### Cell isolation and cryopreservation

Blood collection for peripheral blood mononuclear cells (PBMC) isolation was conducted using KABEVETTE® G EDTA tubes. 2.5 ml of blood was centrifuged at 1500 g for 10 min, the plasma was obtained and stored at -80 °C until further use. The remaining blood was diluted 1:1 with PBS and slowly placed on top of Pancoll human (PAN-Biotech). Density gradient centrifugation was performed at 800 g for 30 min. without break. The interface containing the PBMCs was collected from the gradient and washed twice with PBS at 500 g for 10 minutes. The cell pellet was resuspended in PBS and the cell number was determined in a Sysmex KX-21N (Sysmex Europe GmbH). 1.8×10^7^ cells were cryopreserved in FCS (PAN-Biotech) containing 10% DMSO. The cryotubes were cooled down overnight in a Mr. Frosty (Sigma) at -80 °C and stored in liquid nitrogen until further use.

### Anti-SARS-CoV-2 spike antibody titer

Plasma samples were analyzed for the spike (receptor-binding domain [RBD]; sequence derived from the original wildtype SARS-CoV-2 strain) specific immunoglobulin G antibodies by enzyme-linked immunosorbent assay (ELISA) as described previously ^38^. Briefly, samples were diluted from 1:100 to 1:12,500 and results were expressed as the dilution, which still gave the same signal as an internal calibrator of the ELISA, indicating a positive result. The values obtained for samples below the detection limit were interpreted as negative and set to 1. The assay was calibrated according to the World Health Organization international standards and values were expressed as binding antibody units (BAU).

### SARS-CoV-2 neutralization assay

SARS-CoV-2 pseudoviruses were prepared as described previously ^39^. Briefly, sera were incubated for 30 min at 56°C in order to inactivate complement factors. Single cycle VSV*ΔG(FLuc) pseudoviruses bearing the SARS-CoV-2 spike (D614G) protein ^40^ or SARS-CoV-2 B.1.617.2 (Delta) (EPI_ISL_1921353) spike in the envelope were incubated with quadruplicates of two-fold serial dilutions from 1:20 to 1:2560 of immune sera in 96-well plates prior to infection of Vero E6 cells (1×10^4^ cells/well) in DMEM with 10% FBS (Life Technologies). 18 hours post infection, firefly luciferase (FLuc) reporter activity was determined as previously described ^41^ using a CentroXS LB960 (Berthold). The reciprocal antibody dilution causing 50% inhibition of the luciferase reporter was calculated as pseudovirus neutralization dose 50% (PVND50). Detection range is defined to be between 1:20 and 1:2560.

### PBMC stimulation using SARS-CoV-2 peptide pool

PBMCs were thawed in a 37 °C water bath and diluted in 10 ml RPMI 1640 with Glutamine (Capricorn) with 5 % human AB serum (PAN-Biotech) and 5 U/ml Benzonase (Merck/Sigma) and centrifuged at 400 g for 5 min. The pellet was washed with 10 ml thawing medium, resuspended in 5 ml culture medium (RPMI 1640 with Glutamine (Capricorn) plus 5 % human AB serum (PAN-Biotech)) and cells were rested for at least 2 hours. Cell number was determined using 7-aminoactinomycin D in a Cytoflex LX (Beckman Coulter). 100 µl per sample were seeded in duplicates in a flat bottom 96 well plate (Sarstedt) at a concentration of 1×10^7^ cells/ml. Cells were stimulated with PepTivator pools (Miltenyi Biotec) for 16 hours according to manufacturer’s instructions. The peptide mixes consisted of a pool of mainly 15-mer sequences with 11 amino acids overlap, covering the complete protein coding sequence (aa 5–1273) of the surface or spike glycoprotein (“S”, 130-127-953, Miltenyi Biotec) of SARS Coronavirus 2 (GenBank MN908947.3, Protein QHD43416.1), as well as a mixture of peptides covering the membrane glycoprotein (“M”, 30-126-702, Miltenyi Biotec) and the nucleocapsid phosphoprotein (“N”, 130-126-698, Miltenyi Biotec) protein of SARS-CoV-2. Additional wells included a positive control (CytoStim™, Miltenyi Biotec) and an unstimulated control, in which sterile water instead of the PepTivator pool was added. Except the positive control, all conditions were done in duplicates.

### Flow cytometry

After peptide stimulation for 16 hours, PBMC were stained with the reagents contained in the SARS-CoV-2 T cell analysis kit (PBMC), human plus anti-CD137-PE-Vio 615 (Miltenyi Biotec; see Supplementary Table 1) according to manufacturer’s recommendations and analyzed by flow cytometry.

For analysis of different T cell subsets, 2.5 × 10^6^ PBMCs were stained with viability stain LIVE/DEAD™ Fixable Blue Dead Cell Stain Kit, for UV excitation (L23105, Thermo Fisher) for 30 min. at 4 °C. Afterwards, Fc receptors were blocked by incubating the cells with Human TruStain FcX™ (422302, Biolegend) for 15 min. at 4 °C. Subsequently, surface markers were stained for 15 min. at 4°C. Fixation and permeabilization were performed with Foxp3 staining buffer set (130-093-142, Miltenyi Biotec). Next, intracellular proteins Ki-67 and Foxp3 were stained for 30 min. at 4°C. Antibody dilutions are presented in Supplementary Table 2. Flow cytometry measurement was performed using a Cytoflex LX (Beckman Coulter). Flow cytometry data were analyzed with FlowJo™ (Becton Dickinson & Company, version 10.8.0). Barnes-Hut algorithm was implemented to generate t-Distributed Stochastic Neighbor Embedding (t-SNE) plots with FlowJo™ for high-dimensional data visualization.

### Statistics

Mann-Whitney or Kruskal–Wallis one-way ANOVA tests were performed to calculate statistical significance using Prism (GraphPad Software v9.2.0, San Diego, CA). The data were imported in R program (v3.6.3). Correlation analysis was performed using “rcorr” function of Hmisc package (v4.4-0) and visualized by “ggscatter” function of ggplot2-based ggpubr package (v0.2.5). Spearman method was used to compute the correlation coefficient (R) and p-value ≤ 0.05 was considered as significant correlation. Antibody titers were compared by using the Kruskal–Wallis test with Dunn’s multiple comparison or Mann–Whitney test.

## RESULTS

### Cohort characteristics

The immune response towards COVID-19 vaccines in patients with severe diseases such as cancer has recently been assessed ^42, 43, 44, 45^. However, an in-depth analysis of immune responses in PLWH has not been performed, yet. Therefore, we recruited a cohort of 71 people living with HIV (PLWH) receiving anti-retroviral therapy (ART) (mean age = 46.1 ± 10.9; 62 male and 9 female) and 20 healthy donors (controls; mean age = 39.4 ± 11.9; 8 male and 12 female) to analyze humoral and cellular immune responses after two doses of vaccination. All participants received the BNT162b2 vaccine (Pfizer-BioNTech). Except three participants, all of the PLWH cohort had a CD4 count higher than 200 cells/μl before and 3 months after vaccination (mean CD4 count: 756.4 cells/mm^3^; range: 79 – 1562 cells/mm^3^). None in the PLWH cohort reported a confirmed SARS-CoV-2 infection. One participant of the healthy control group had a confirmed SARS-CoV-2 infection and got a BNT162b2 vaccination six months after the infection as recommended by the German Robert Koch Institute (RKI, Berlin). Since the responses of this donor were in the range of other healthy donors, we did not exclude these values. HIV viral load was >30 copies/ml (range: 58-1573 copies/ml) in 6 participants at time of first vaccination. All but one (191 copies/ml) declined during follow up below detection level. The clinical data are summarized in Table 1. Blood samples were drawn immediately before the first (T0) and second (T1) vaccination and four to six weeks after the second vaccination (T2) (Fig. 1A).

**Fig. 1.**
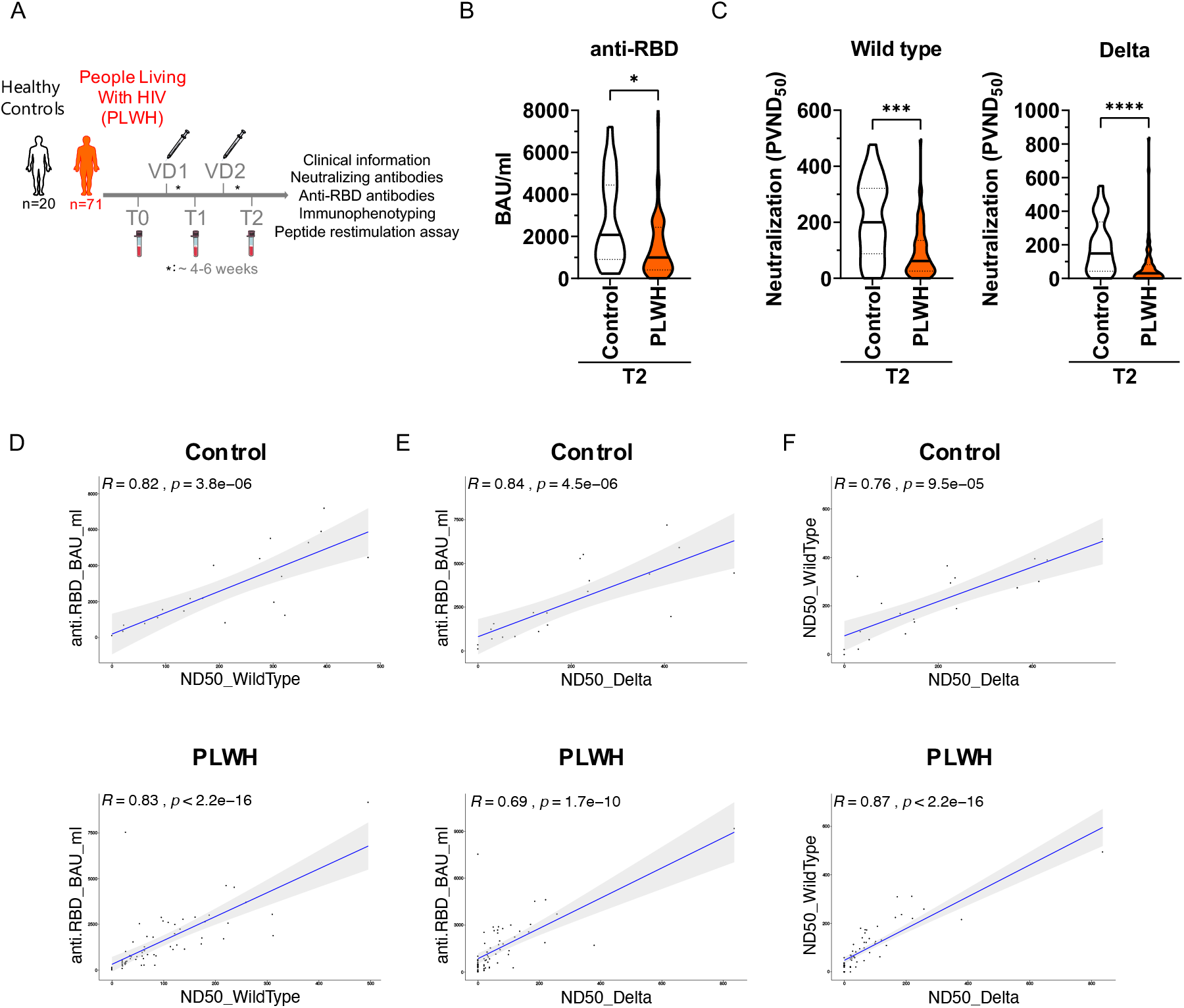
Humoral immune response in vaccinated PLWH and healthy controls. **(A)** Scheme showing the two groups of the study - control (n=20) and people living with HIV (PLWH; n= 71) - and the time points of blood sampling (T0, T1, T2) and vaccination shots (V1, V2). **(B)** anti-RBD antibodies were determined by ELISA. Values are given as Binding Antibody Units (BAU) per ml. (PLWH T0, n= 71; PLWH T1, n= 61; PLWH T2, n= 68; Control T2, n= 20). **(C)** Neutralizing antibodies to SARS-CoV-2 Wuhan (PLWH T2, n= 70; Control T2, n= 20) and Delta (PLWH T2, n= 65; Control T2, n= 20) strains. Vero E6 cells were infected with the respective pseudoviruses in the presence of serial dilutions of sera. The pseudotype virus neutralization dose 50% (PVND50) is plotted. **(D-F)** The scatter plots represent correlation analysis between indicated parameters for control and PLWH groups. Black dots are the individual human donors. The regression line is indicated in blue, while gray area is 95% confidence interval region. In the graphs, R is abbreviated for correlation coefficient and p stands for p-value of statistical significance. Statistical significance was calculated by two-tailed Mann-Whitney test: *p < 0.05, ***p < 0.001, ****p < 0.0001.

### Reduced humoral immune responses in PLWH upon COVID-19 vaccination

During SARS-CoV-2 infection, most of the antibody response is directed against the receptor-binding domain (RBD) of the SARS-CoV-2 spike protein and serum levels of anti-RBD antibodies correlate well with the humoral immune response upon vaccination as well as protection against SARS-CoV-2 ^6, 46^. Therefore, we quantified anti-RBD antibodies in the plasma of study participants by ELISA. Antibody titers after two doses of vaccine administration were reduced (GMT: 838.42 BAU/ml; 95% confidence interval: 430.79-1246.05) compared to healthy control (GMT: 1841.94 BAU/ml; 95% confidence interval: 850.35-2833.53) (Fig. 1B). Nevertheless, vaccination induced anti-RBD antibody titers in PLWH and the second dose of vaccination increased further these titers significantly (Supplementary Fig. 1). Notably, two donors in the PLWH group had detectable antibody titers already at T0 suggesting that they had a previous asymptomatic SARS-CoV-2 infection (Supplementary Fig. 1).

Next, we analyzed the capacity of sera to neutralize SARS-CoV-2 infection. To this end, we employed an established pseudovirus assay ^39^ and determined the neutralization dose against the viral strain first reported in Wuhan, China (referred to hereafter as WT) and Delta SARS-CoV-2 strains. The amounts of neutralizing antibodies were highly reduced in PLWH compared to healthy controls for both, WT (p = 0.0004) and Delta (p <0.0001) SARS-CoV-2 strains (Fig. 1C). Nearly the same statistically significant correlation was found between anti-RBD antibody response and neutralization capacity against WT SARS-CoV-2 strain in healthy participants (R=0.82, p=3.8e-06) as well as PLWH cohorts (R=0.83, p<2.2e-16) (Fig. 1D). However, the correlation analyses for anti-RBD antibody and neutralization against SARS-CoV-2 Delta strain were remarkably decreased in PLWH patients (R=0.69, p=1.7e-10) and healthy controls (R=0.84, p=4.5e-06) (Fig. 1E). The neutralization capacity against WT and Delta strains of SARS-CoV-2 correlated better in PLWH patients (R=0.87, p<2.2e-16) compared to healthy individuals (R=0.76, p=9.5e-05) (Fig. 1F), which might be due to the higher number of participants in the PLWH cohort. We conclude that although SARS-CoV-2 vaccination induces antibody responses in PLWH, but the extent of the humoral immune response is reduced compared to healthy controls.

### Altered cytotoxic T cell subsets and constitutive activation of CD8+ T cells in PLWH

Next, we analyzed cellular immune responses towards SARS-CoV-2 vaccination in PLWH and control groups. Here, multi-parameter flow cytometry was employed and the gating strategy is shown in Supplementary Fig. 2. We started with analyzing the frequencies of T cells and the two major T cell lineages, CD4+ helper and CD8+ cytotoxic cells. We detected no differences in the frequencies of total T cells as marked by CD3 expression between PLWH and the control group (Fig. 2A). As expected, the frequency of CD4+ T cells was reduced in PLWH compared to the controls (Fig. 2B), while the frequency of CD8+ T cells was increased (Fig. 2C). Accordingly, the ratio of CD4 to CD8 T cells was significantly lower in PLWH compared to the control group (Fig. 2D). Of note, the frequency of PD-1 expressing CD4+ T cells, a marker associated with an exhausted phenotype, was increased in PLWH compared to controls (Fig. 2E). The frequency of CD19+ B cells was reduced in PLWH compared to the control group (Fig. 2F).

**Fig. 2.**
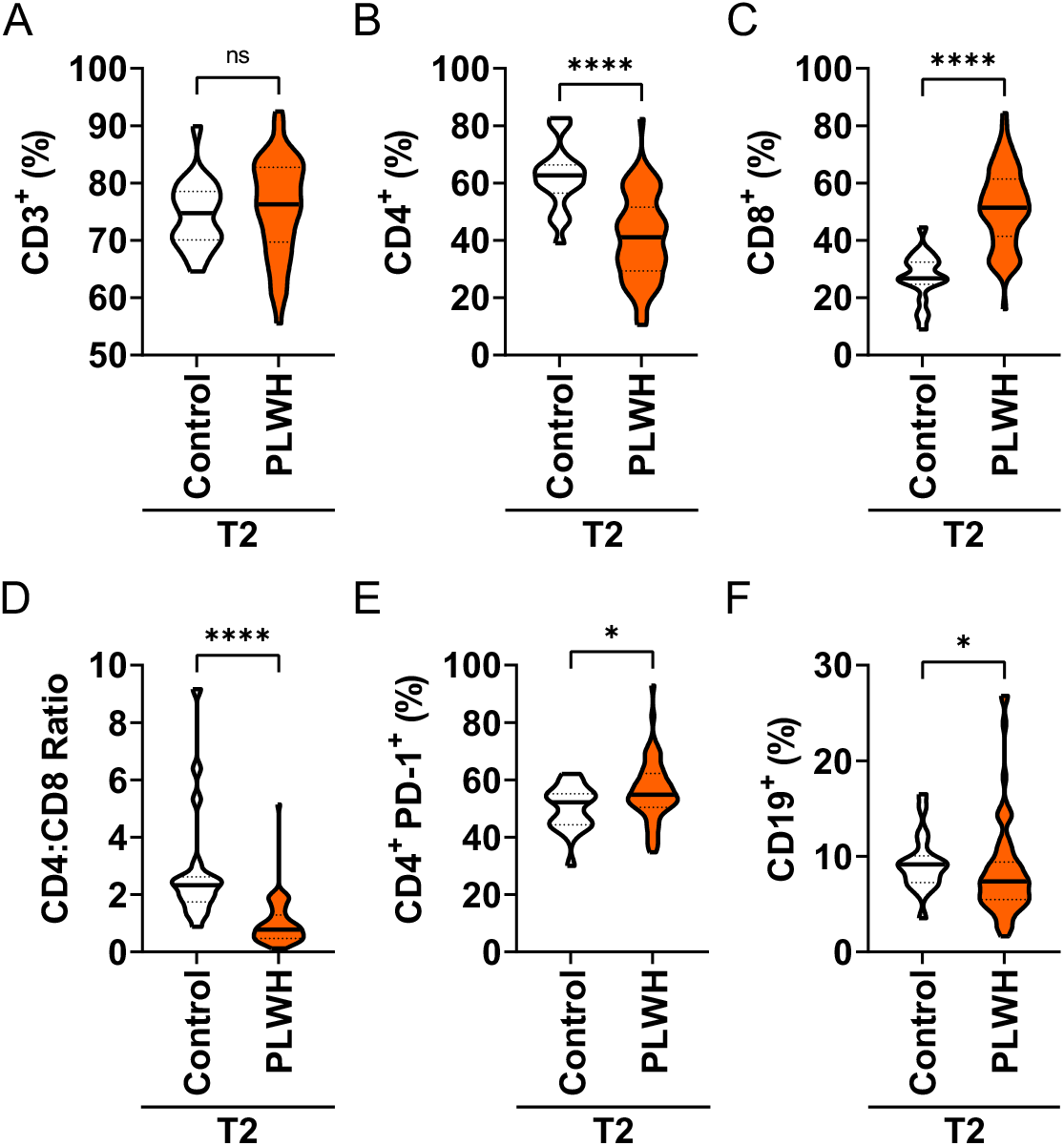
Frequency of lymphocyte lineages in vaccinated PLWH and healthy controls. **(A)** The frequency of CD3+ T cells after two doses of vaccine (T2) is plotted for the control and PLWH groups. **(B)** Frequency of CD4+ T cells. **(C)** Frequency of CD8+ T cells. **(D)** The CD4:CD8 ratio is plotted for the control and PLWH groups. **(E)** The frequency of PD-1+ cells within the CD4+ compartment is plotted for the control and PLWH groups. **(F)** Frequency of CD19+ B cells in controls and PLWH at time point T2. Statistical significance was calculated by two-tailed Mann-Whitney test: *p < 0.05, ****p < 0.0001 (PLWH T0, n= 65; PLWH T1, n= 54; PLWH T2, n= 64; Control T2, n= 20).

Within the CD8+ compartment, the naïve CD8+ T cells (CD27+ CD45RA+ CCR7+) were reduced in PLWH compared to controls (Fig. 3A). In line, naïve CD8+ T cells of PLWH exhibited decreased proliferation as analyzed by the expression of Ki-67 (Fig. 3B). Moreover, we detected increased frequencies of effector (E; CD27-CD45RA+ CCR7-; Fig. 3C) and effector memory (EM; CD27-CD45RA-CCR7-; Fig. 3D) CD8+ T as well as decreased frequencies of transitional memory (TM; CD27+ CD45RA-CCR7-; Fig. 3E) CD8+ T cells in PLWH. The observation that CD8+ T cells are less naïve and more in an activated and effector-like state in PLWH than in healthy controls was also supported by unsupervised clustering using t-SNE for two representative donors (Fig. 3F). Thus, the phenotypic profiling of the CD8 T cell compartment suggests a high constitutive activation of these cells. In line with this notion, we detected increased frequencies of activated, i.e. CD137 expressing, CD8+ cells that co-expressed the inflammatory cytokines IFNγ and TNFα even in the absence of any antigenic peptide stimulation making it impossible to measure antigen-specific responses in the CD8 compartment (Supplementary Fig. 3A-C). Moreover, increased frequencies of CD8+ CD137+ IFNγ+ TNFα+ T cells were detected in PLWH upon stimulation with peptide pools derived from spike (S) protein as well as membrane plus nucleocapsid proteins (M+N) (Supplementary Fig. 3B-E) suggesting that this response is unlikely to be related to the vaccination. Taken together, similar to what has been reported for SARS-CoV-2 infection in PLWH ^37^, we found that ART did not fully reconstitute the T cell compartment in PLWH and that CD8+ T cells exhibit an activated phenotype.

**Fig. 3.**
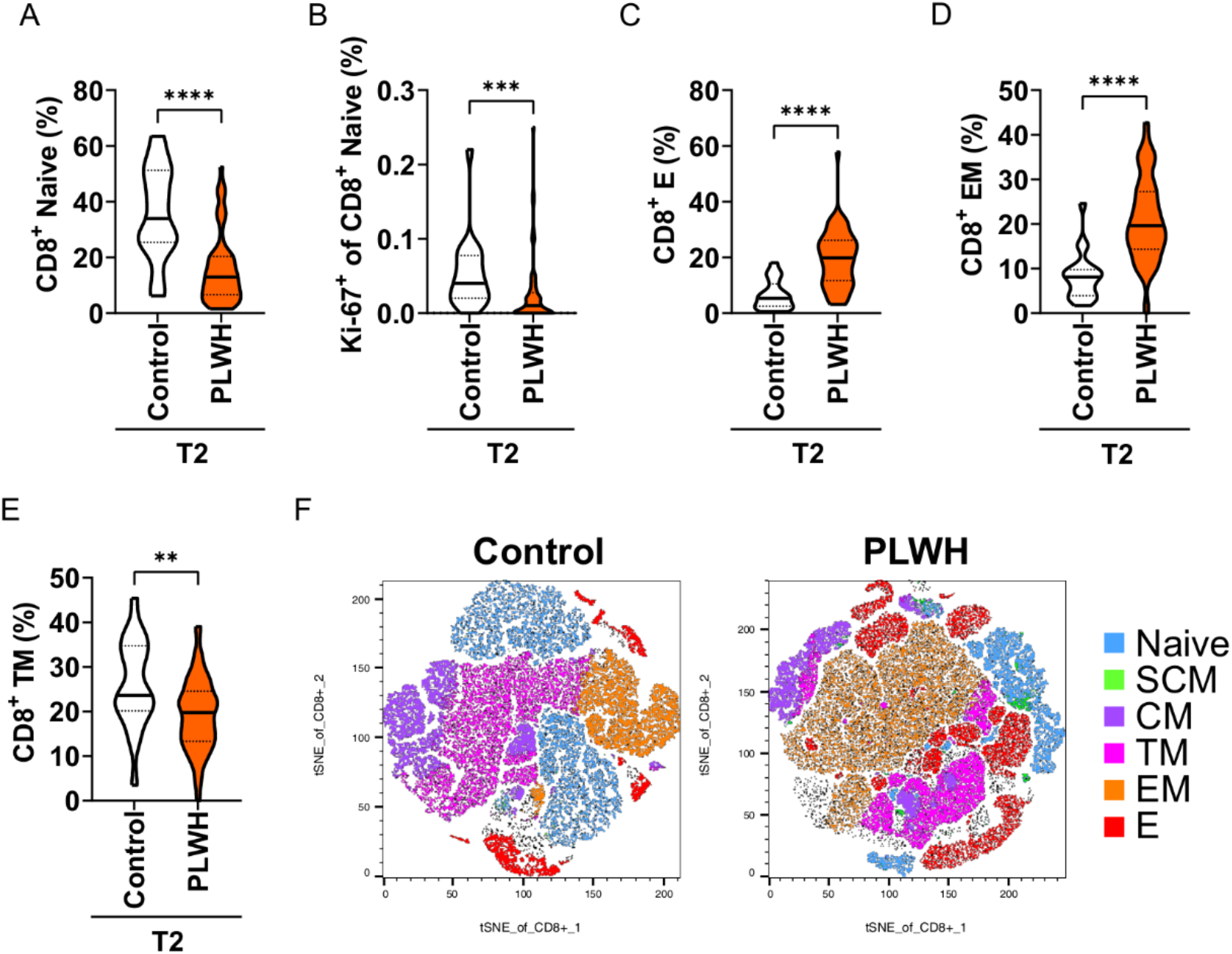
Phenotype and antigen-dependent responses of CD8+ cytotoxic T cells in vaccinated PLWH and healthy controls. **(A)** Frequency of naïve CD8+ T cells. **(B)** Frequency of proliferating (Ki-67 positive) naïve CD8+ T cells. **(C)** Frequency of effector CD8+ T cells. **(D)** Frequency of effector memory CD8+ T cells. **(E)** Frequency of transitional memory CD8+ T cells. **(F)** t-SNE plots of the distribution of naïve, stem cell-like memory (SCM), central memory (CM), transitional memory (TM), effector memory (EM) and effector (E) CD8+ T cells for control and PLWH groups. Statistical significance was calculated by two-tailed Mann-Whitney test: **p < 0.01, ***p < 0.001, ****p < 0.0001 (PLWH T0, n= 65; PLWH T1, n= 54; PLWH T2, n= 64; Control T2, n= 20).

### Comparable CD4+ T cells responses in PLWH and healthy controls upon vaccination

Furthermore, we analyzed cellular responses of CD4+ T helper cells upon vaccination. Surprisingly, we detected decreasing frequencies of circulating follicular T helper (cTFH) cells, which are characterized by the expression of CXCR5 and PD-1, over the course of vaccination in PLWH (Fig. 4A). Consistent with the situation observed for CD8+ T cells in PLWH, we found increased frequencies of CD25+ activated conventional T (Tcon) cells and reduced frequencies of naïve Tcon cells after full vaccination with two doses (Fig. 4B-C). Similarly, the CD4+ compartment of PLWH contained higher frequencies of terminally differentiated CD4+ T cells (TD; CD27-CD45RA+ CCR7-; Fig. 4D), and of effector memory T cells (EM; CD27-CD45RA-CCR7-; Fig. 4E). Again, unsupervised clustering using t-SNE of two representative donors supported our findings that CD4+ T cells of PLWH are rather in an activated and differentiated state and that the naïve phenotype is reduced (Fig. 4F).

**Fig. 4.**
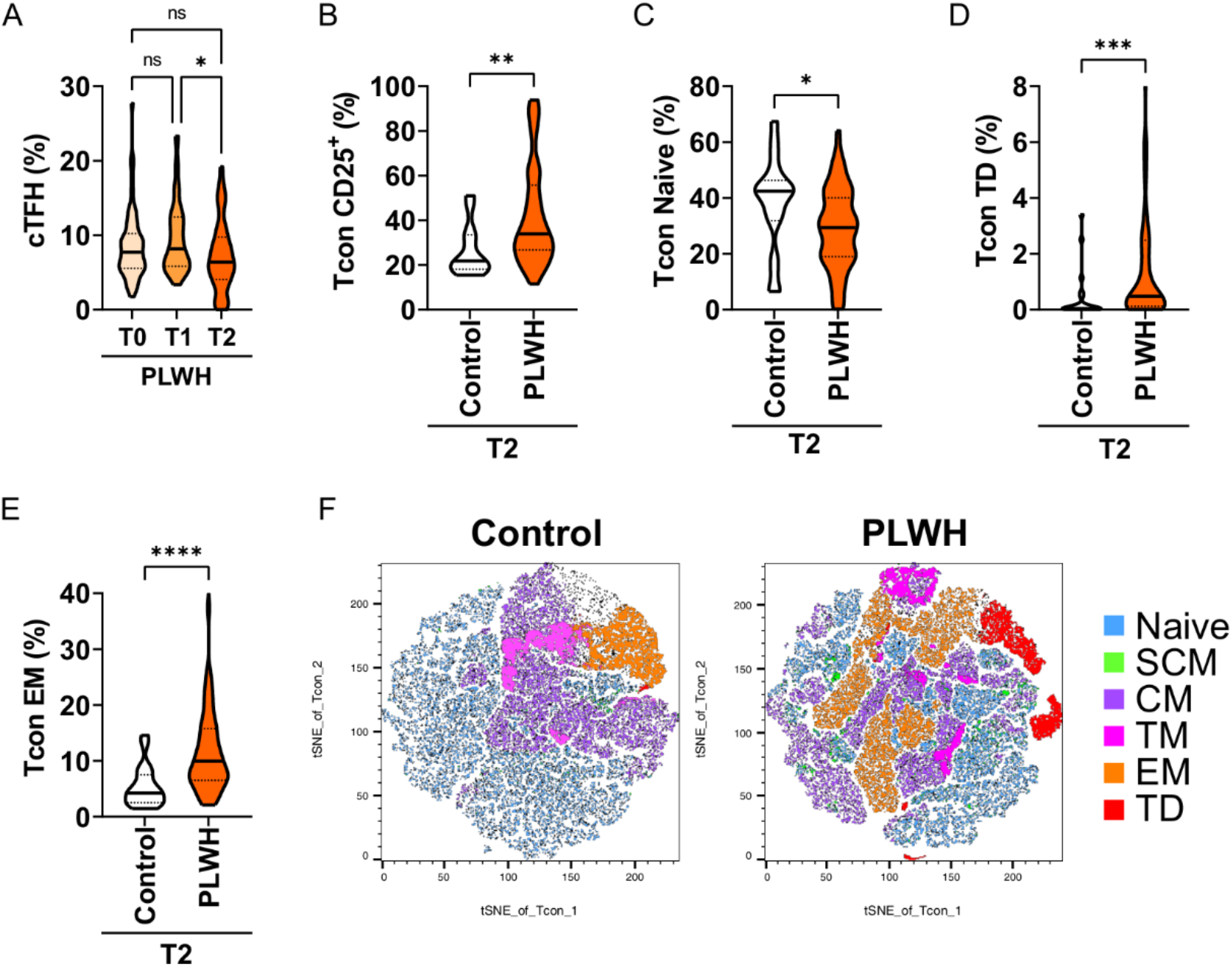
Immune phenotyping of CD4+ helper T cells in vaccinated PLWH and healthy controls. **(A)** Frequency of circulating follicular CD4+ T helper cells in PLWH before the first vaccination (T0), before the second vaccination (T1) and 4 to 6 weeks after the second vaccination (T2). **(B)** Frequency of activated (CD25 positive) conventional CD4+ T cells at T2 in control and PLWH groups. **(C)** Frequency of naive conventional CD4+ T cells. **(D)** Frequency of terminally differentiated conventional CD4+ T cells. **(E)** Frequency of effector memory conventional CD4+ T cells. **(F)** t-SNE plots of the distribution of naïve, stem cell-like memory (SCM), central memory (CM), transitional memory (TM), effector memory (EM) and terminally differentiated (TD) CD4+ T cells for control and PLWH groups. Statistical significance was calculated by Kruskal-Wallis test and Dunn’s test for multiple comparisons (a) or two-tailed Mann-Whitney test (b-e): **p < 0.01, ***p < 0.001, ****p < 0.0001 (PLWH T0, n= 65; PLWH T1, n= 54; PLWH T2, n= 64; Control T2, n= 20).

Finally, we analyzed antigen-specific responses in CD4+ T helper cells. We detected only minimal cytokine responses in cultures without any peptide stimulation or in cultures stimulated with peptide pools derived from membrane (M) and nucleocapsid (N) proteins (Fig. 5A and Supplementary Fig. 4A). In contrast, polyclonal stimulation (positive control (Pos) with CytoStim™) resulted in strong responses at all time points (Fig. 5A). Stimulation with spike (S) protein peptide pools resulted in upregulation of the activation markers CD137 and CD154 on CD4+ T cells of PLWH dependent on the number of vaccination doses (Fig. 5B). We found a significant anti-correlation between S-peptide antigen-specific CD4 T cells expressing activation markers CD154+CD137+ and the ratio of CD4 to CD8 in the PLWH cohort (R=-0.39; p-value=0.0033; Fig. 5C lower scatter plot), which was insignificant in healthy controls (R=-0.31; p-value=0.18; Fig. 5C upper scatter plot). Thus, low CD4+ relative to CD8+ T cell counts are surprisingly associated with better activation of CD4+ T cells.

**Fig. 5.**
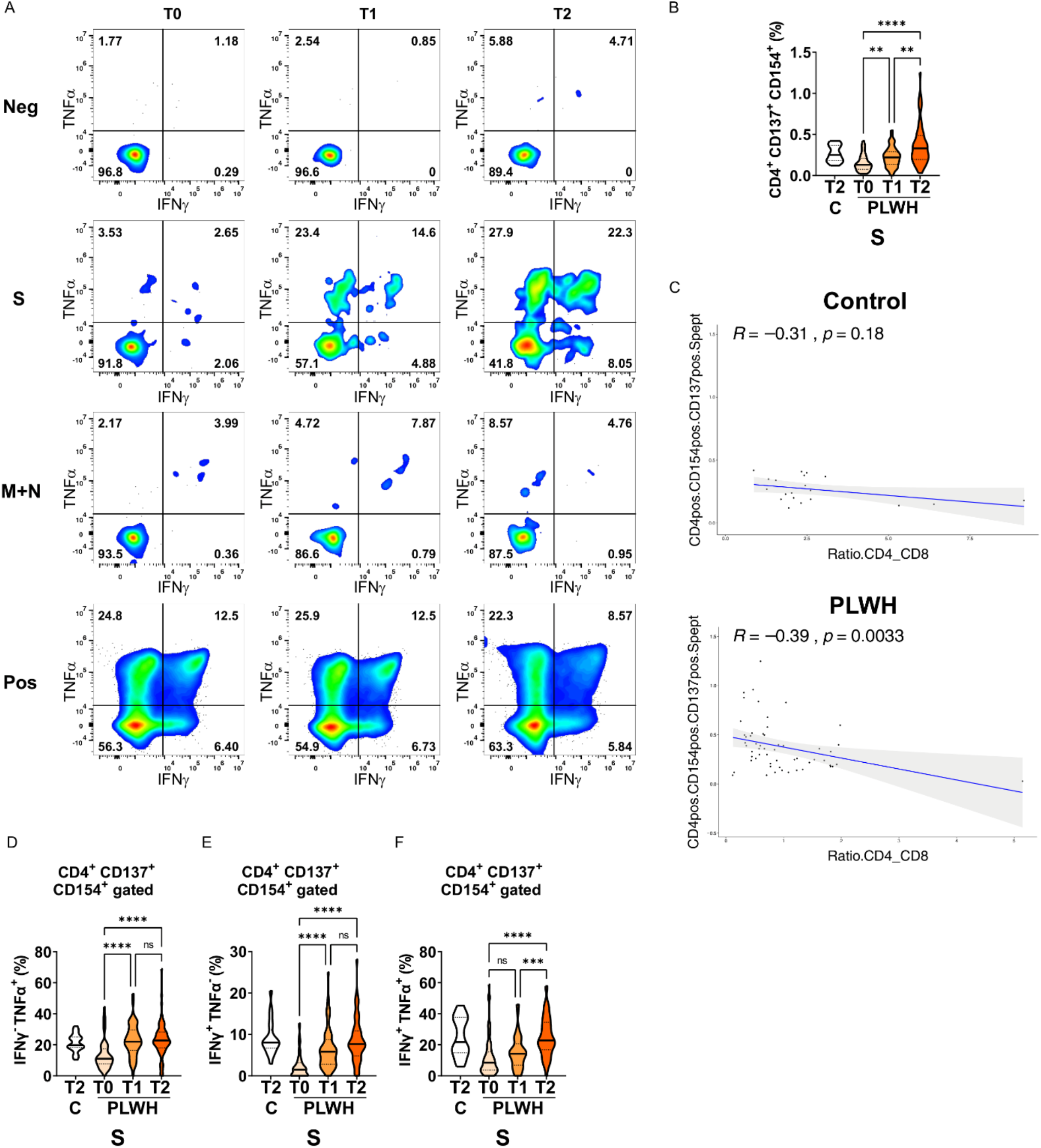
Antigen-dependent responses of CD4+ helper T cells in vaccinated PLWH and healthy controls. **(A)** Representative dot plots of TNFα and IFNγ expression in activated CD4+ CD137+ CD154+ T cells of a PLWH donor stimulated with SARS-CoV-2 spike (S) glycoprotein peptide pool, or with membrane glycoprotein + nucleocapsid phosphoprotein (M+N) peptide pool, or with CytoStim™ (Pos), or left untreated (Neg) before and after vaccination shots. **(B)** The violin plot represents frequency of activated, i.e. CD137+ CD154+, CD4+ T cells and **(C)** their correlation (spearman method) with CD4:CD8 ratio from healthy control subjects (upper scatter plot) and PLWH (lower scatter plot). The violin plots represent **(D)** frequency of TNFα+ cells in activated CD4+ CD137+ CD154+ T cells, **(E)** frequency of IFNγ+ cells in activated CD4+ CD137+ CD154+ T cells and **(F)** frequency of TNFα+ IFNγ+ double-positive cells in activated CD4+ CD137+ CD154+ T cells. Statistical significance was calculated by Kruskal-Wallis test and Dunn’s test for multiple comparisons: **p < 0.01, ***p < 0.001, ****p < 0.0001 (PLWH T0, n= 64-65; PLWH T1, n= 54; PLWH T2, n= 59-61; Control T2, n= 20). Abbreviations used: C, control group; PLWH, people living with HIV; T0, before first vaccination; T1, before second vaccination; T2, 4 to 6 weeks after second vaccination; S, spike protein-derived peptide pool.

We then measured IFNγ and TNFα expression in the CD154 and CD137 co-expressing CD4+ T cells and found a robust increase in single (IFNγ or TNFα) or double (IFNγ and TNFα) cytokine-producing cells in PLWH after the first and the second vaccination (Fig. 5D-F). Importantly, the expression of activation markers and cytokines at the T2 time point exhibited no significant differences between PLWH and the control group. Therefore, despite the known reduced frequency of CD4+ T helper cells in PLWH, the antigen-specific responses in the CD4 compartment are comparable between PLWH and control groups.

The correlation analysis at the T2 time point in the healthy control participants showed a significant positive correlation between S-peptide antigen-specific CD4+CD154+CD137+ T cells producing IFNγ+TNFα+ cells and neutralizing capacity against SARS-CoV-2 WT strain (R=0.54; p-value=0.015; Fig. 6A left graph), delta variant (R=0.64, p-value=0.0022; Fig. 6B left graph) and with anti-RBD titer (R=0.66; p-value=0.0016; Fig. 6C left graph). Interestingly, the correlation value R of these comparisons in PLWH cohort exhibited a significant weaker association (Fig. 6A-C right graphs) and a non-significant correlation for anti-RBD (R=0.18; p-value=0.18). Furthermore, comparable findings were obtained when activated CD4+ T cells that only expressed the activation marker CD154 were analyzed for S-peptide stimulation (Supplementary Fig. 4B-D) and correlated with neutralization against delta variant (Supplementary Fig. 4E). In conclusion, while the cellular immune response by CD4+ T cells is surprisingly comparable to the control group, the humoral response mediated by antibodies is reduced suggesting that PLWH might be less protected from COVID-19 by vaccination.

**Fig. 6.**
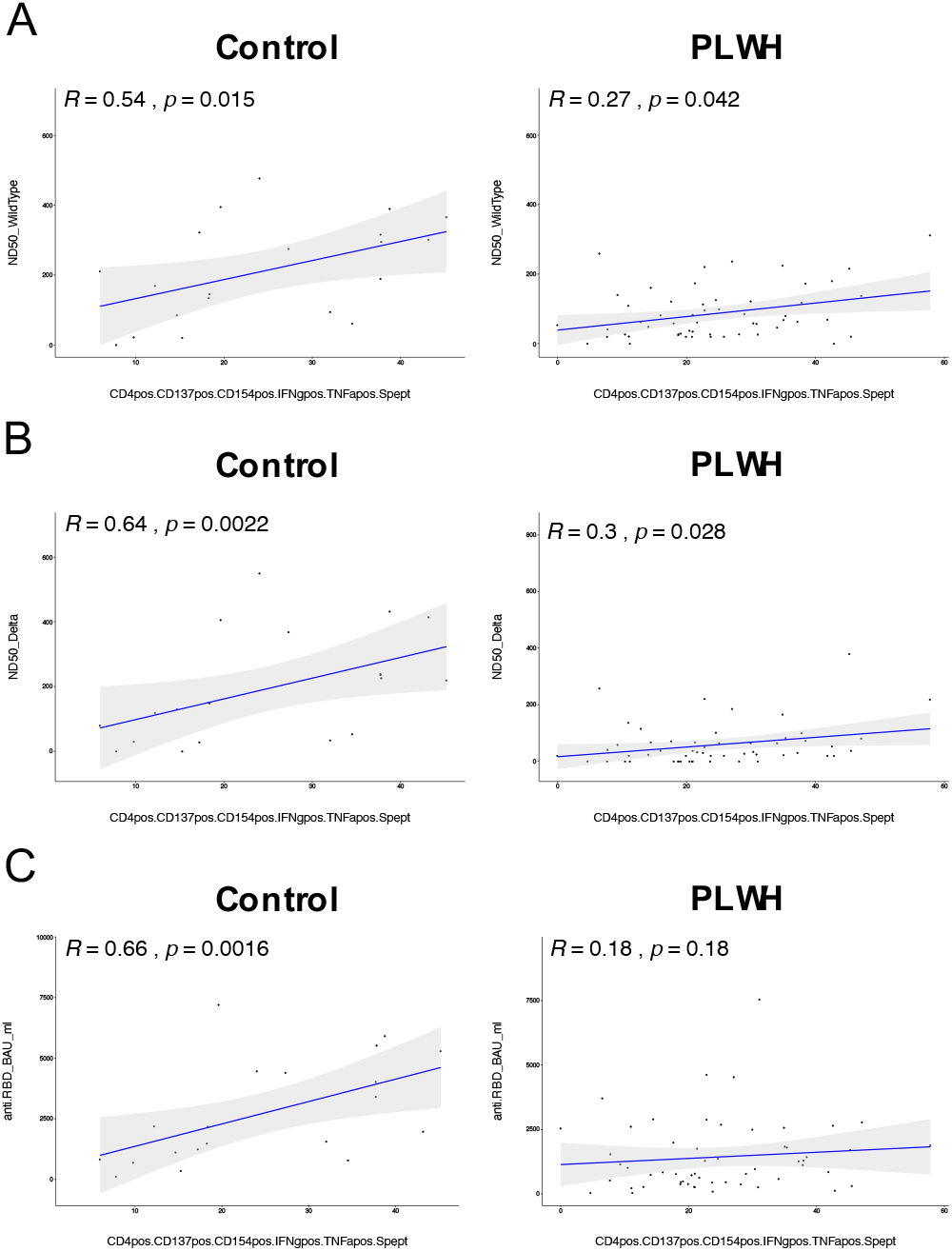
Correlation analysis of cytokine responses in activated T cells and humoral response in vaccinated PLWH and healthy controls. CD4+ CD137+ CD154+ T cells expressing both IFNγ+ and TNFα+ were correlated using spearman method for T2 i.e. after two doses of vaccination. The scatter plots represent correlation between CD4+ CD137+ CD154+ IFNγ+ TNFα+ T cells and neutralizing capacity against WT strain of SARS-CoV-2 **(A)**, delta variant of SARS-CoV-2 **(B)** and against anti-RBD titers **(C)** on the left side for healthy controls, while scatter plots on the right side are for PLWH cohort. The blue line indicates a regression line and gray area shows 95% confidence interval. Abbreviations used: R, correlation coefficient; p, p-value for the statistical significance.

## DISCUSSION

The occurrence of SARS-CoV-2 in the human population in late 2019 led to a pandemic that is causing a major health care burden worldwide. In an effort to cope with the pandemic, several vaccines have been developed that reduce the risk to suffer from severe COVID-19 and of mortality by inducing humoral and cellular immune responses (reviewed in ^47^). Particularly the novel mRNA vaccines show high efficacy and are well tolerated ^16, 17, 48^. However, COVID-19 vaccines may induce less efficient immune responses in the context of immunosuppression. In this line, antibody responses to spike protein in people with inborn errors of immunity were reduced compared to controls when CD3+ T cells counts were lower than 1000 cells/ml or CD19+ B cell counts were lower than 100 cells/ml ^49^. Moreover, impaired antibody, B cell and T cell responses have been reported for patients receiving kidney transplantation receiving COVID-19 mRNA vaccination ^50, 51, 52, 53, 54^.

Here, we addressed the question whether COVID-19 mRNA vaccines are able to mount immune responses in PLWH, since HIV can cause acquired immunodeficiency. To do so, we analyzed humoral, i.e. antibody, and cellular immune responses in PLWH over the time course of vaccination and compared the responses after two doses of vaccine with healthy controls, who were fully vaccinated. We detected anti-RBD antibodies and neutralizing antibodies against the original strain (WT) of SARS-CoV-2 as well as against the Delta variant in PLWH albeit at lower levels than in the control group. Especially the neutralizing antibody titers against the Delta variant were low in PLWH and the general antibody titers correlated less well with neutralizing antibodies against Delta as compared to the correlation of anti-RBD antibodies versus neutralizing antibodies against the SARS-CoV-2 WT strain. This might indicate an improved immune evasion of the Delta variant and raises concerns about the protection that two doses of mRNA vaccine mount in PLWH against current and upcoming variants of concern of SARS-CoV-2. Our results on the humoral immune response are in agreement with other reports that reported on immunogenicity and safety of COVID-19 vaccines ^55, 56, 57^. In contrast, similar antibody responses were reported in a PLWH cohort that received an inactivated SARS-CoV-2 vaccine ^58^.

Next to antibody titers, we also analyzed cellular immune responses in PLWH upon COVID-19 vaccination by multi-parametric flow cytometry. As expected ^59^, we detected a reduced CD4+ to CD8+ T cell ratio and CD8+ T cells exhibited a more activated phenotype. A reduced naïve CD8 compartment and a higher prevalence of effector, effector memory and transitional memory CD8+ T cells was not only observed by conventional flow cytometrical analysis but also by unsupervised t-SNE clustering. Additionally, the activated phenotype of CD8+ T cells prevented the analysis of antigen-specific CD8+ T cell responses since these cells expressed the type I immunity signature cytokines IFNγ and TNFα already in the absence of any stimulation in our in vitro cultures.

A higher activation and a reduced naïve compartment in PLWH were also observed for CD4+ T cells, again by conventional flow cytometrical analysis and unsupervised clustering using t-SNE. Nevertheless, we were able to analyze antigen-specific CD4+ T cell responses since the expression of cytokines in the absence of any re-stimulating peptides was negligible. Of note, we detected higher responses to the peptide pool containing M and N derived peptides in PLWH after full vaccination as compared to the control group (Supplementary Fig. 4A). One explanation could be that a few donors of the PLWH cohort contracted a SARS-CoV-2 infection. Although no case of symptomatic COVID-19 was reported to us during sampling of the PLWH cohort, we indeed detected anti-RBD antibodies in two PLWH donors suggesting asymptomatic SARS-CoV-2 infections. An alternative explanation might be that these responses are due to previous exposures to other human corona viruses, which can cause common cold symptoms and might share common epitopes with SARS-CoV-2. More importantly, we detected an increase in the expression of the activation marker CD137 and CD154 as well as in the expression of the cytokines IFNγ and TNFα by CD4+ T cells over the time course of vaccination. Notably, the expression of these markers and the frequency of multifunctional T cells, which express both IFNγ and TNFα, was similar in PLWH and in the control group. Thus, despite the reduced CD4+ T cell count in the peripheral blood of PLWH, the CD4+ cellular response to COVID-19 vaccination is preserved. Furthermore, we found a positive correlation between the frequency of multifunctional CD4+ T cells (CD4+ CD137+ CD154+ IFNγ+ and TNFα+) and neutralizing antibody responses in PLWH. However, the correlation was less pronounced in comparison to the one in healthy controls, and the correlation was lost for multifunctional CD4+ T cells and anti-RBD antibodies.

In addition, we found that the CD4 to CD8 ratio negatively correlated with T cell activation, as detected by the presence of CD4+ CD137+ CD154+ cells, in response to antigens derived from the spike protein in PLWH. No such significant correlation was found in the healthy control group. Therefore, a reduction in CD4+ relative to CD8+ T cells appears to result in better T cell activation in PLWH. Moreover, the CD4 to CD8 ratio might be a predictive biomarker for the effectiveness of COVID-19 vaccines in PLWH. Similar observations have been made for hepatitis B virus and Yellow Fever virus vaccines ^60, 61^.

The diminished antibody responses detected in our PLWH cohort are in line with reports of reduced antibody titers upon vaccination of PLWH with vaccines against influenza A virus ^62, 63, 64^ or BCG ^65^. However, while some of these studies also reported reduced T cell responses ^62, 63^, we found similar CD4+ T cell responses in PLWH vaccinated with an COVID-19 mRNA vaccine. Whether these differences are due to differences in the viral antigen or due to the different vaccine formulation remains to be tested in future studies.

While the CD4+ T cell responses towards spike protein-derived peptides are promising, the reduced antibody responses raise the question whether two doses of mRNA vaccine are sufficient to protect PLWH from COVID-19. Recent evidence suggests that a booster by a third dose of vaccine not only enhances antibody levels, which might decrease over time, but also broadens the antibody-mediated immunity and can protect against variants of concern ^66, 67, 68, 69^. Therefore, additional booster vaccinations might be required for PLWH to maintain or to reach full protection from COVID-19.

### Limitations of the study

The PLWH participants in our study received well-adjusted anti-retroviral therapy (ART) and had CD4+ T cells counts higher than 200 cells/μl. Whether immune responses upon COVID-19 vaccination correlate with T cell numbers in the peripheral blood awaits the analysis of a larger cohort with more diverse T cell counts. Moreover, we did not test directly for vaccine safety or efficacy since this would be beyond the scope of this study. Nevertheless, no gross adverse effects were reported by the participants of our PLWH cohort. Furthermore, our results suggest that vaccination of PLWH with COVID-19 mRNA vaccines might elicit partial humoral and cellular immune protection. Future studies have to show, whether or not booster vaccination will enhance immune protection, especially against upcoming variants of concern.

## Data Availability

All data produced in the present study are available upon reasonable request to the authors.

## Acknowledgments

We would like to thank all blood donors who participated in this study. We are grateful to all members of the WIR - Walk In Ruhr, Center for Sexual Health and Medicine, Bochum, Germany, for supporting this study. We thank Gert Zimmer, Institute for Virology und Immunology, Switzerland and Department of Infectious Diseases and Pathobiology (DIP), Vetsuisse Faculty, University of Bern, Switzerland as well as Stefan Pöhlmann and Markus Hoffmann, Infection Biology Unit, German Primate Center - Leibniz Institute for Primate Research, Göttingen, Germany, Faculty of Biology and Psychology, Georg-August-University Göttingen, Göttingen, Germany, for providing the WT und Delta plasmid.

## Author contributions

AP, ASR and NHB acquired patient samples; CB, CPS, CM, EVB, PB, SB, SS and DU performed experiments; CB, CPS, JB, SP and IS analyzed data; CB, CPS, JB and IS wrote the draft of the manuscript; all authors read and approved the final version of the manuscript; AP, SF, OO, ARS, CW, SP and IS designed the study; AP and IS supervised the study.

## References

1. Zhou, P. et al. A pneumonia outbreak associated with a new coronavirus of probable bat origin. Nature 579, 270–273 (2020).

2. Shields, A.M., Burns, S.O., Savic, S., Richter, A.G. & Consortium, U.P.C.-. COVID-19 in patients with primary and secondary immunodeficiency: The United Kingdom experience. J Allergy Clin Immunol 147, 870–875 e871 (2021).

3. Williamson, E.J. et al. Factors associated with COVID-19-related death using OpenSAFELY. Nature 584, 430–436 (2020).

4. Breton, G. et al. Persistent cellular immunity to SARS-CoV-2 infection. J Exp Med 218 (2021).

5. Grifoni, A. et al. Targets of T Cell Responses to SARS-CoV-2 Coronavirus in Humans with COVID-19 Disease and Unexposed Individuals. Cell 181, 1489–1501 e1415 (2020).

6. Ju, B. et al. Human neutralizing antibodies elicited by SARS-CoV-2 infection. Nature 584, 115–119 (2020).

7. Ni, L. et al. Detection of SARS-CoV-2-Specific Humoral and Cellular Immunity in COVID-19 Convalescent Individuals. Immunity 52, 971–977 e973 (2020).

8. Peng, Y. et al. Broad and strong memory CD4(+) and CD8(+) T cells induced by SARS-CoV-2 in UK convalescent individuals following COVID-19. Nat Immunol 21, 1336–1345 (2020).

9. Robbiani, D.F. et al. Convergent antibody responses to SARS-CoV-2 in convalescent individuals. Nature 584, 437–442 (2020).

10. Suthar, M.S. et al. Rapid Generation of Neutralizing Antibody Responses in COVID-19 Patients. Cell Rep Med 1, 100040 (2020).

11. Dagan, N. et al. BNT162b2 mRNA Covid-19 Vaccine in a Nationwide Mass Vaccination Setting. N Engl J Med 384, 1412–1423 (2021).

12. Chaudhary, N., Weissman, D. & Whitehead, K.A. mRNA vaccines for infectious diseases: principles, delivery and clinical translation. Nat Rev Drug Discov 20, 817–838 (2021).

13. Shang, J. et al. Cell entry mechanisms of SARS-CoV-2. Proc Natl Acad Sci U S A 117, 11727–11734 (2020).

14. Anderson, E.J. et al. Safety and Immunogenicity of SARS-CoV-2 mRNA-1273 Vaccine in Older Adults. N Engl J Med 383, 2427–2438 (2020).

15. Dan, J.M. et al. Immunological memory to SARS-CoV-2 assessed for up to 8 months after infection. Science 371 (2021).

16. Polack, F.P. et al. Safety and Efficacy of the BNT162b2 mRNA Covid-19 Vaccine. N Engl J Med 383, 2603–2615 (2020).

17. Sahin, U. et al. COVID-19 vaccine BNT162b1 elicits human antibody and TH1 T cell responses. Nature 586, 594–599 (2020).

18. Widge, A.T. et al. Durability of Responses after SARS-CoV-2 mRNA-1273 Vaccination. N Engl J Med 384, 80–82 (2021).

19. Guerrera, G. et al. BNT162b2 vaccination induces durable SARS-CoV-2-specific T cells with a stem cell memory phenotype. Sci Immunol 6, eabl5344 (2021).

20. Turner, J.S. et al. SARS-CoV-2 mRNA vaccines induce persistent human germinal centre responses. Nature 596, 109–113 (2021).

21. Kustin, T. et al. Evidence for increased breakthrough rates of SARS-CoV-2 variants of concern in BNT162b2-mRNA-vaccinated individuals. Nat Med 27, 1379–1384 (2021).

22. Levin, E.G. et al. Waning Immune Humoral Response to BNT162b2 Covid-19 Vaccine over 6 Months. N Engl J Med 385, e84 (2021).

23. Thompson, M.G. et al. Effectiveness of a Third Dose of mRNA Vaccines Against COVID-19-Associated Emergency Department and Urgent Care Encounters and Hospitalizations Among Adults During Periods of Delta and Omicron Variant Predominance - VISION Network, 10 States, August 2021-January 2022. MMWR Morb Mortal Wkly Rep 71, 139–145 (2022).

24. Byrd, K.M. et al. SARS-CoV-2 and HIV coinfection: clinical experience from Rhode Island, United States. J Int AIDS Soc 23, e25573 (2020).

25. Harter, G. et al. COVID-19 in people living with human immunodeficiency virus: a case series of 33 patients. Infection 48, 681–686 (2020).

26. Karmen-Tuohy, S. et al. Outcomes Among HIV-Positive Patients Hospitalized With COVID-19. J Acquir Immune Defic Syndr 85, 6–10 (2020).

27. Shalev, N. et al. Clinical Characteristics and Outcomes in People Living With Human Immunodeficiency Virus Hospitalized for Coronavirus Disease 2019. Clin Infect Dis 71, 2294–2297 (2020).

28. Sigel, K. et al. Coronavirus 2019 and People Living With Human Immunodeficiency Virus: Outcomes for Hospitalized Patients in New York City. Clin Infect Dis 71, 2933–2938 (2020).

29. Stoeckle, K. et al. COVID-19 in Hospitalized Adults With HIV. Open Forum Infect Dis 7, ofaa327 (2020).

30. Vizcarra, P. et al. Description of COVID-19 in HIV-infected individuals: a single-centre, prospective cohort. Lancet HIV 7, e554–e564 (2020).

31. Bhaskaran, K. et al. HIV infection and COVID-19 death: a population-based cohort analysis of UK primary care data and linked national death registrations within the OpenSAFELY platform. Lancet HIV 8, e24–e32 (2021).

32. Dandachi, D. et al. Characteristics, Comorbidities, and Outcomes in a Multicenter Registry of Patients With Human Immunodeficiency Virus and Coronavirus Disease 2019. Clin Infect Dis 73, e1964–e1972 (2021).

33. Geretti, A.M. et al. Outcomes of Coronavirus Disease 2019 (COVID-19) Related Hospitalization Among People With Human Immunodeficiency Virus (HIV) in the ISARIC World Health Organization (WHO) Clinical Characterization Protocol (UK): A Prospective Observational Study. Clin Infect Dis 73, e2095–e2106 (2021).

34. Hoffmann, C. et al. Immune deficiency is a risk factor for severe COVID-19 in people living with HIV. HIV Med 22, 372–378 (2021).

35. Tesoriero, J.M. et al. COVID-19 Outcomes Among Persons Living With or Without Diagnosed HIV Infection in New York State. JAMA Netw Open 4, e2037069 (2021).

36. Spinelli, M.A. et al. SARS-CoV-2 seroprevalence, and IgG concentration and pseudovirus neutralising antibody titres after infection, compared by HIV status: a matched case-control observational study. Lancet HIV 8, e334–e341 (2021).

37. Alrubayyi, A. et al. Characterization of humoral and SARS-CoV-2 specific T cell responses in people living with HIV. Nat Commun 12, 5839 (2021).

38. Urlaub, D., Wolfsdorff, N., Durak, D., Renken, F. & Watzl, C. SARS-CoV-2 infection shortly after BNT162b2 vaccination results in high anti-spike antibody levels in nursing home residents and staff. Immun Inflamm Dis 9, 1702–1706 (2021).

39. Zettl, F. et al. Rapid Quantification of SARS-CoV-2-Neutralizing Antibodies Using Propagation-Defective Vesicular Stomatitis Virus Pseudotypes. Vaccines (Basel) 8 (2020).

40. Hoffmann, M. et al. SARS-CoV-2 Cell Entry Depends on ACE2 and TMPRSS2 and Is Blocked by a Clinically Proven Protease Inhibitor. Cell 181, 271–280 e278 (2020).

41. Kinast, V. et al. C19orf66 is an interferon-induced inhibitor of HCV replication that restricts formation of the viral replication organelle. J Hepatol 73, 549–558 (2020).

42. Ehmsen, S. et al. Antibody and T cell immune responses following mRNA COVID-19 vaccination in patients with cancer. Cancer Cell 39, 1034–1036 (2021).

43. Greenberger, L.M. et al. Antibody response to SARS-CoV-2 vaccines in patients with hematologic malignancies. Cancer Cell 39, 1031–1033 (2021).

44. Monin, L. et al. Safety and immunogenicity of one versus two doses of the COVID-19 vaccine BNT162b2 for patients with cancer: interim analysis of a prospective observational study. Lancet Oncol 22, 765–778 (2021).

45. Van Oekelen, O. et al. Highly variable SARS-CoV-2 spike antibody responses to two doses of COVID-19 RNA vaccination in patients with multiple myeloma. Cancer Cell 39, 1028–1030 (2021).

46. Gilbert, P.B. et al. Immune correlates analysis of the mRNA-1273 COVID-19 vaccine efficacy clinical trial. Science 375, 43–50 (2022).

47. Sadarangani, M., Marchant, A. & Kollmann, T.R. Immunological mechanisms of vaccine-induced protection against COVID-19 in humans. Nat Rev Immunol 21, 475–484 (2021).

48. Baden, L.R. et al. Efficacy and Safety of the mRNA-1273 SARS-CoV-2 Vaccine. N Engl J Med 384, 403–416 (2021).

49. Delmonte, O.M. et al. Antibody responses to the SARS-CoV-2 vaccine in individuals with various inborn errors of immunity. J Allergy Clin Immunol 148, 1192–1197 (2021).

50. Havlin, J. et al. Immunogenicity of BNT162b2 mRNA COVID-19 vaccine and SARS-CoV-2 infection in lung transplant recipients. J Heart Lung Transplant 40, 754–758 (2021).

51. Miele, M. et al. Impaired anti-SARS-CoV-2 humoral and cellular immune response induced by Pfizer-BioNTech BNT162b2 mRNA vaccine in solid organ transplanted patients. Am J Transplant 21, 2919–2921 (2021).

52. Rincon-Arevalo, H. et al. Impaired humoral immunity to SARS-CoV-2 BNT162b2 vaccine in kidney transplant recipients and dialysis patients. Sci Immunol 6 (2021).

53. Sattler, A. et al. Impaired humoral and cellular immunity after SARS-CoV-2 BNT162b2 (tozinameran) prime-boost vaccination in kidney transplant recipients. J Clin Invest 131 (2021).

54. Stumpf, J. et al. Humoral and cellular immunity to SARS-CoV-2 vaccination in renal transplant versus dialysis patients: A prospective, multicenter observational study using mRNA-1273 or BNT162b2 mRNA vaccine. Lancet Reg Health Eur 9, 100178 (2021).

55. Feng, Y. et al. Safety and immunogenicity of inactivated SARS-CoV-2 vaccine in high-risk occupational population: a randomized, parallel, controlled clinical trial. Infect Dis Poverty 10, 138 (2021).

56. Levy, I. et al. Immunogenicity and safety of the BNT162b2 mRNA COVID-19 vaccine in people living with HIV-1. Clin Microbiol Infect 27, 1851–1855 (2021).

57. Milano, E. et al. Immunogenicity and safety of the BNT162b2 COVID-19 mRNA vaccine in PLWH: A monocentric study in Bari, Italy. J Med Virol (2022).

58. Liu, Y. et al. COVID-19 Vaccination in People Living with HIV (PLWH) in China: A Cross Sectional Study of Vaccine Hesitancy, Safety, and Immunogenicity. Vaccines (Basel) 9 (2021).

59. Warren, J.A., Clutton, G. & Goonetilleke, N. Harnessing CD8(+) T Cells Under HIV Antiretroviral Therapy. Front Immunol 10, 291 (2019).

60. Avelino-Silva, V.I. et al. CD4/CD8 Ratio Predicts Yellow Fever Vaccine-Induced Antibody Titers in Virologically Suppressed HIV-Infected Patients. J Acquir Immune Defic Syndr 71, 189–195 (2016).

61. Fuster, F. et al. CD4/CD8 ratio as a predictor of the response to HBV vaccination in HIV-positive patients: A prospective cohort study. Vaccine 34, 1889–1895 (2016).

62. George, V.K. et al. HIV infection Worsens Age-Associated Defects in Antibody Responses to Influenza Vaccine. J Infect Dis 211, 1959–1968 (2015).

63. Parmigiani, A. et al. Impaired antibody response to influenza vaccine in HIV-infected and uninfected aging women is associated with immune activation and inflammation. PLoS One 8, e79816 (2013).

64. Tebas, P. et al. Poor immunogenicity of the H1N1 2009 vaccine in well controlled HIV-infected individuals. AIDS 24, 2187–2192 (2010).

65. Mansoor, N. et al. HIV-1 infection in infants severely impairs the immune response induced by Bacille Calmette-Guerin vaccine. J Infect Dis 199, 982–990 (2009).

66. Choi, A. et al. Safety and immunogenicity of SARS-CoV-2 variant mRNA vaccine boosters in healthy adults: an interim analysis. Nat Med 27, 2025–2031 (2021).

67. Garcia-Beltran, W.F. et al. mRNA-based COVID-19 vaccine boosters induce neutralizing immunity against SARS-CoV-2 Omicron variant. Cell 185, 457–466 e454 (2022).

68. Levine-Tiefenbrun, M. et al. Viral loads of Delta-variant SARS-CoV-2 breakthrough infections after vaccination and booster with BNT162b2. Nat Med 27, 2108–2110 (2021).

69. Shroff, R.T. et al. Immune responses to two and three doses of the BNT162b2 mRNA vaccine in adults with solid tumors. Nat Med 27, 2002–2011 (2021).

