## Supplementary material and figures for "Impact of SARS-CoV-2 vaccination on systemic immune responses in people living with HIV"

### Supplementary Materials and Methods

### Antibody panels used for flow cytometry

**Supplementary Table 1: PepTivator Assay**

| Specificity | Clone | Fluorochrome | Dilution |
|---|---|---|---|
| CD3 | REA613 | APC | 1:50 |
| CD4 | REA623 | VioBright B515 | 1:50 |
| CD8 | REA734 | VioGreen | 1:50 |
| CD14 | REA599 | VioBlue | 1:50 |
| CD20 | REA780 | VioBlue | 1:50 |
| CD137 (4-1BB) | REA765 | PE-Vio 615 | 1:50 |
| CD154 (CD40L) | REA238 | APC-Vio 770 | 1:50 |
| IFN-γ | REA600 | PE | 1:50 |
| TNF-α | REA656 | PE-Vio 770 | 1:50 |
| Viobility Fixable Dye | - | 405/452 | 1:100 |

**Supplementary Table 2**

| Specificity | Clone | Fluorochrome | Cat. number | Company | Dilution |
|---|---|---|---|---|---|
| CD3 | UCHT1 | Brilliant Violet 510 | 300448 | Biolegend | 1:100 |
| CD4 | OKT4 | PerCP-Cy5.5 | 344608 | Biolegend | 1:500 |
| CD8 | RPA-T8 | APC- Fire 750 | 344746 | Biolegend | 1:500 |
| CD19 | HIB19 | AlexaFluor 700 | 302225 | Biolegend | 1:500 |
| CD25 | BC96 | Brilliant Violet 421 | 302630 | Biolegend | 1:200 |
| CD27 | O323 | Brilliant Violet 785 | 302831 | Biolegend | 1:100 |
| CD45RA | HI100 | PerCP | 304155 | Biolegend | 1:100 |
| CD95 | DX2 | Brilliant Violet 650 | 305642 | Biolegend | 1:100 |

| | | | | | |
|---|---|---|---|---|---|
| CD107a | H4A3 | Brilliant Violet 711 | 328639 | Biolegend | 1:200 |
| CD127 | HIL-7R-M21 | BUV737 | 612794 | BD Biosciences | 1:20 |
| CD185 (CXCR5) | J252D4 | AlexaFluor 488 | 356911 | Biolegend | 1:100 |
| CD197 (CCR7) | G043H7 | APC | 353213 | Biolegend | 1:100 |
| CD1279 (PD-1) | EH12.2H7 | PE-Dazzle 594 | 329939 | Biolegend | 1:20 |
| Foxp3 | 206D | PE | 320107 | Biolegend | 1:20 |
| Ki-67 | Ki-67 | PE-Cy7 | 350525 | Biolegend | 1:1000 |

**Supplementary Figures**

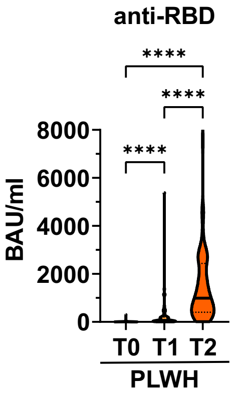


**Supplementary Figure 1** Titers of anti-RBD antibodies in PLWH and PT groups. Statistical significance was calculated by two-tailed Mann-Whitney test: ****p < 0.0001 (PLWH T0, n= 71; PLWH T1, n= 61; PLWH T2, n= 68; Control T2, n= 20).

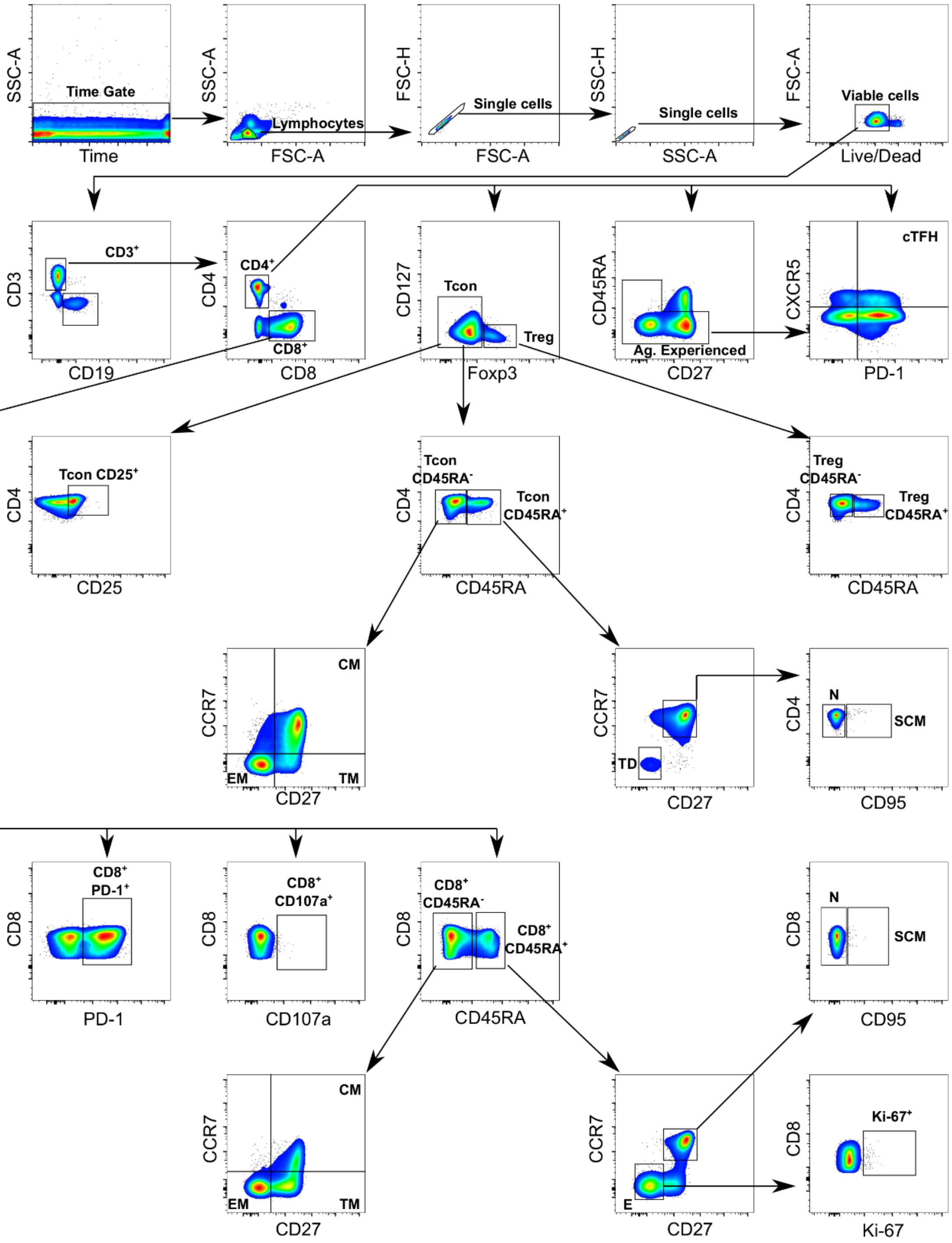
SSC-A
Time Gate
Time
SSC-A
Lymphocytes
FSC-A
FSC-H
Single cells
FSC-A
SSC-H
Single cells
SSC-A
FSC-A
Viable cells
Live/Dead
CD3
CD3+
CD19
CD4
CD4+
CD8+
CD8
CD127
Tcon
Treg
Foxp3
CD45RA
Ag. Experienced
CD27
CXCR5
cTFH
PD-1
CD4
Tcon CD25+
CD25
Tcon CD45RA-
Tcon CD45RA+
CD4
CD45RA
Treg CD45RA-
Treg CD45RA+
CD4
CD45RA
CCR7
CM
EM
TM
CD27
CCR7
TD
CD27
CD4
N
SCM
CD95
CD8
CD8+ PD-1+
PD-1
CD8
CD8+ CD107a+
CD107a
CD8
CD8+ CD45RA-
CD8+ CD45RA+
CD45RA
CD8
N
SCM
CD95
CCR7
CM
EM
TM
CD27
CCR7
E
CD27
CD8
Ki-67+
Ki-67

**Supplementary Figure 2** Gating strategy for analysis of T cell subsets via flow cytometry. Abbreviations used: cTFH (circulating follicular T helper cells), N (Naïve), SCM (Stem cell-like memory), CM (Central memory), TM (Transitional memory), EM (Effector memory), E (Effector), TD (Terminally differentiated).

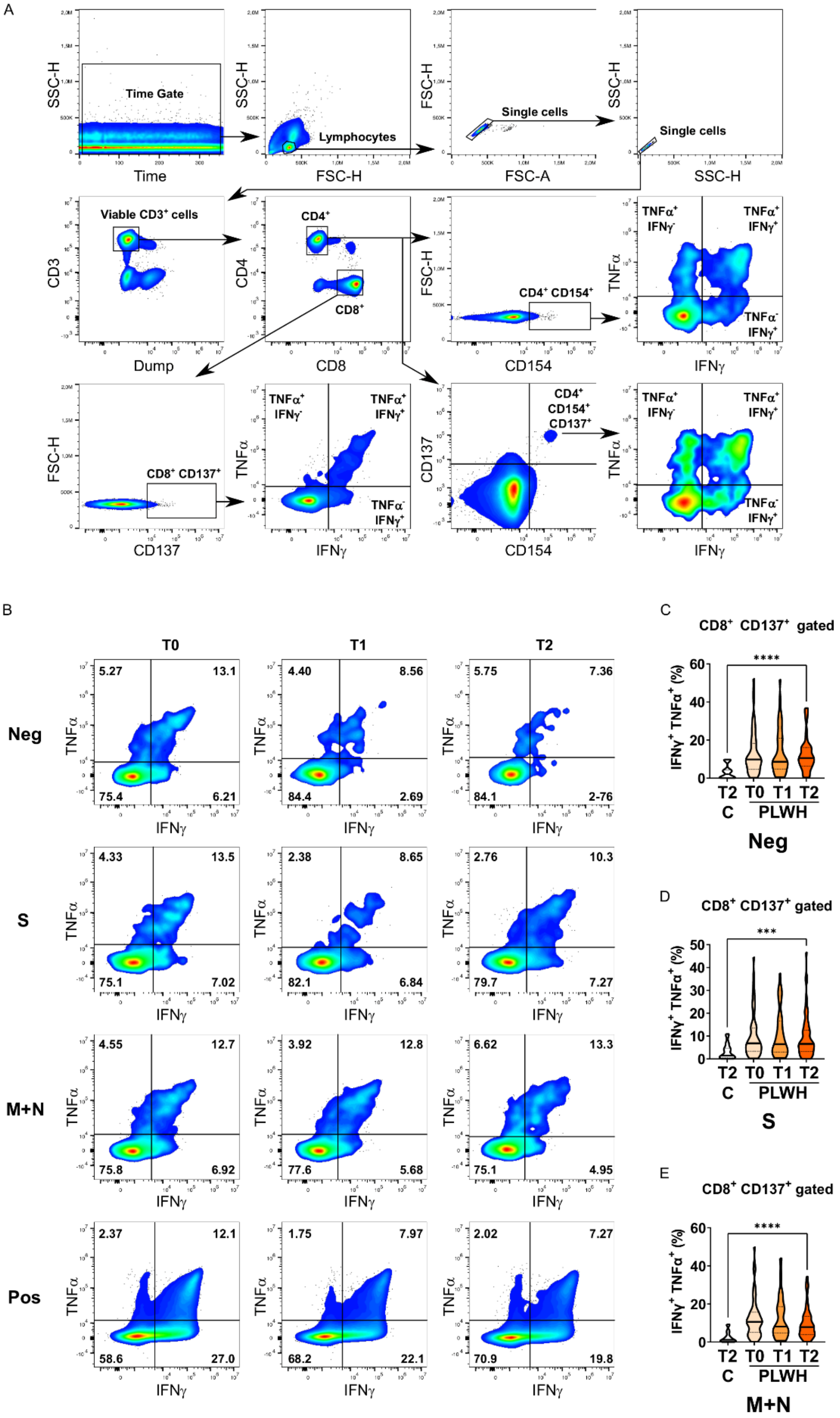
A
Time Gate
SSC-H
Time
Lymphocytes
FSC-H
Single cells
FSC-A
Single cells
SSC-H
Viable CD3+ cells
CD3
Dump
CD4+
CD8+
CD4
CD8
CD4+ CD154+
FSC-H
CD154
TNFα+ IFNγ-
TNFα+ IFNγ+
TNFα- IFNγ+
TNFα
IFNγ
CD8+ CD137+
FSC-H
CD137
CD4+ CD154+ CD137+
CD137
CD154
B
T0
T1
T2
Neg
5.27
13.1
75.4
6.21
4.40
8.56
84.4
2.69
5.75
7.36
84.1
2-76
S
4.33
13.5
75.1
7.02
2.38
8.65
82.1
6.84
2.76
10.3
79.7
7.27
M+N
4.55
12.7
75.8
6.92
3.92
12.8
77.6
5.68
6.62
13.3
75.1
4.95
Pos
2.37
12.1
58.6
27.0
1.75
7.97
68.2
22.1
2.02
7.27
70.9
19.8
C
CD8+ CD137+ gated
****
IFNγ+ TNFα+ (%)
T2 T0 T1 T2
C PLWH
Neg
D
CD8+ CD137+ gated
***
S
E
CD8+ CD137+ gated
****
M+N

**Supplementary Figure 3** Antigen-specific responses of CD8+ cytotoxic T cells. **(A)** Gating strategy for analysis of antigen-specific T cells via flow cytometry. **(B)** Representative dot plots of TNFα and IFNγ expression in activated CD8+ CD137+ CD154+ T cells of a PLWH donor stimulated with SARS-CoV-2 spike (S) glycoprotein peptide pool, or with membrane glycoprotein + nucleocapsid phosphoprotein (M+N) peptide pool, or with CytoStim™ (Pos), or left untreated (Neg) before and after vaccination shots. The violin plots represent **(C)** frequency of activated, i.e. CD137+, CD8+ T cells that produce the inflammatory cytokines IFNγ and TNFα without any peptide stimulation (negative control) in controls and PLWH, **(D)** frequency of activated, i.e. CD137+, CD8+ T cells that produce the inflammatory cytokines IFNγ and TNFα upon stimulation with SARS-CoV-2 spike protein-derived peptides (S) in controls and PLWH and **(E)** frequency of activated, i.e. CD137+, CD8+ T cells that produce the inflammatory cytokines IFNγ and TNFα upon stimulation with peptides derived from SARS-CoV-2 membrane and nucleocapsid proteins that are conserved between different corona viruses (M+N) in controls and PLWH. Statistical significance was calculated by two-tailed Mann-Whitney test: ***p < 0.001, ****p < 0.0001 (PLWH T0, n= 64-65; PLWH T1, n= 54; PLWH T2, n= 59-61; Control T2, n= 20).

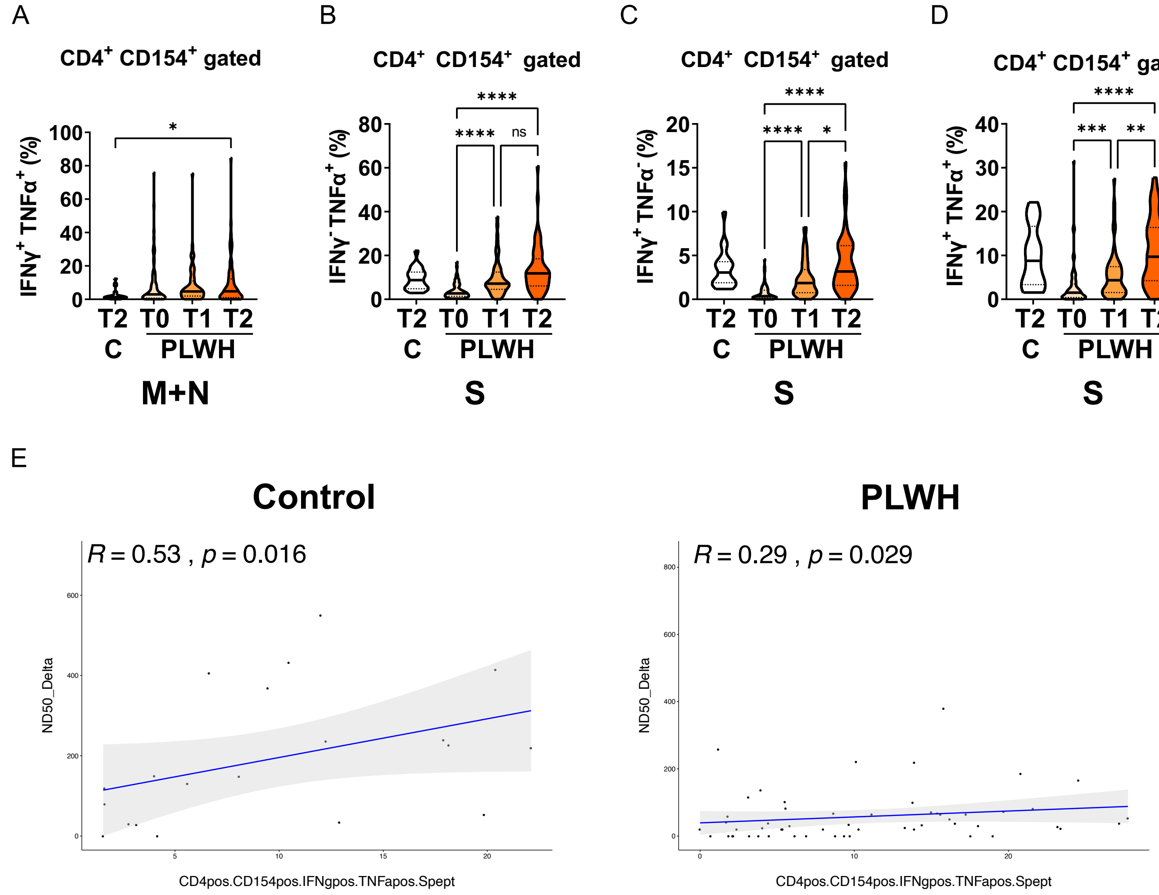


**Supplementary Figure 4** Antigen-specific responses of CD4+ helper T cells. **(A)** Frequency of activated, i.e. CD154+, CD4+ T cells that produce the inflammatory cytokines IFNg and TNFa upon stimulation with peptides derived from SARS-CoV-2 membrane and nucleocapsid proteins that are conserved between different corona viruses (M+N) in controls and PLWH. **(B)** Frequency of activated CD4+ T cells expressing CD154 that produce the inflammatory cytokine TNFα upon stimulation with SARS-CoV-2 spike protein-derived peptides (S) in controls and PLWH. **(C)** Frequency of activated CD4+ T cells expressing CD154 that produce the inflammatory cytokine IFNγ upon stimulation with SARS-CoV-2 spike protein-derived peptides (S) in controls and PLWH. **(D)** Frequency of activated CD4+ T cells expressing CD154 that produce the inflammatory cytokines IFNγ and TNFα upon stimulation with SARS-CoV-2 spike protein-derived peptides (S) in controls and PLWH. Statistical significance was calculated by two-tailed Mann-Whitney test: *$p < 0.05$, **$p < 0.01$, ***$p < 0.001$, ****$p < 0.0001$ (PLWH T0, n= 64-65; PLWH T1, n= 54; PLWH T2, n= 59-61; Control T2, n= 20). **(E)** The spearman's correlation between CD4+ CD154+ IFNγ+ TNFα+ T cells and neutralizing titers against delta variant of SARS-CoV-2 are represented in the scatter plot for healthy participants (left side) and PLWH cohort (right side). R stands for correlation coefficient and p stands for p-value of statistical significance.
